# Pharmacogenomics implementation training improve self-efficacy and competency to drive adoption in clinical practice

**DOI:** 10.1101/2020.11.25.20233858

**Authors:** Fadhli Adesta, Caroline Mahendra, Kathleen Irena Junusmin, Arya Melissa Selva Rajah, Sharon Goh, Levana Sani, Alexandre Chan, Astrid Irwanto

**Author notes:** These authors contributed equally to this work (Fadhli Adesta, Caroline Mahendra).

## Abstract

**Background:** Administration of pharmacogenomics (PGx) testing in clinical practice has been suboptimal, presumably due to lack of PGx education. Here, we aim to evaluate the standpoint of PGx testing among a diverse group of healthcare professionals (HCPs) through conducting surveys before and after training.

**Materials & Methods:** Training modules were designed to cover 3 key learning objectives and deployed in 5 sections. A pre- and post-training survey questionnaire was used to evaluate participants self-assessments on employing PGx in clinical practice.

**Results & Conclusion:** Out of all enrollments, 102 survey responses were collected. Overall, respondents agree on the benefits of PGx testing, but have inadequate self-efficacy and competency in utilizing PGx data. Our results show that training significantly improve these, and even leading to greater anticipation of PGx adoption.

## INTRODUCTION

Pharmacogenomics (PGx) focuses on the influence of genetic variations on drug response [1]. PGx is progressing from identifying drug-gene pairs to assimilating into clinical practice [2]. A recent study conducted in Singapore observed that 30% of adverse drug reactions (ADRs) were caused by at least one drug with a PGx clinical annotation, suggesting the potential to prevent ADR occurrence via PGx testing [3]. Despite PGx testing demonstrating its potential to enhance medication safety and efficacy [4], its utility in clinical practice has been suboptimal [5], specifically in Asia [6]. The lack of PGx education is an often-cited barrier to the widespread implementation of PGx [7–10].

Although PGx didactic teaching is increasing in undergraduate and postgraduate schools of medicine and pharmacy [2,10–15], PGx education is not readily available to practising clinicians [5,10,16]. Clinicians may not have sufficient training and educational background to offer patient care incorporating PGx and personalized care overall [5,8]. Consequently, their poor perceived ability to clinically integrate PGx has been widely reported [5,9,10,16,17]. In particular, a survey on Singaporean clinicians practising in psychiatry observed that only 46.4% of respondents felt competent to order PGx tests [5]. In this regard, PGx education may help to bridge the knowledge translation gap of PGx use among clinicians [16].

PGx educational courses may be the key to encouraging greater assimilation of PGx into routine practice, having proven to improve attitudes [7] and increase the adoption of testing [18]. A study conducted on physicians observed that a 45-minute PGx presentation can improve their attitudes towards PGx testing [7]. PGx educational courses would have to be constantly updated to ensure sustainable PGx assimilation into routine clinical practice [4,6]. This study is novel because this is the first training material that considers the current level of understanding in Asian healthcare professionals towards pharmacogenomics. It is also novel because of the evaluation of such training material being tested to offline and online healthcare professionals. Therefore, this study aims to evaluate the development and outcomes of a PGx implementation training programme. Participants’ perceptions of the clinical relevance and utility of PGx, and their self-efficacy and knowledge to integrate PGx into practice were assessed.

## MATERIALS AND METHODS

### Study Design and Subjects

This is a mixed method study incorporating two phases: (1) development of training materials and (2) training evaluation measures. In the first phase (development phase), we build the training materials consisting of two versions, TM1 (prototype) and TM2 (finalized). The flow of training delivery is as stated in Fig 1b. Both trainings were conducted by licensed pharmacists who are certified in PGx through American Society of Health-System Pharmacists (ASHP). In the second phase, we assess the designed training materials through pre- and post-surveys after each training. Healthcare professionals (HCPs) were recruited to participate in the focus group discussion (FGD) including medical practitioners (both general practitioners and specialists) as well as pharmacists and students for diversity. Written consent was obtained from FGD participants, highlighting voluntary participation.

**Fig. 1.**
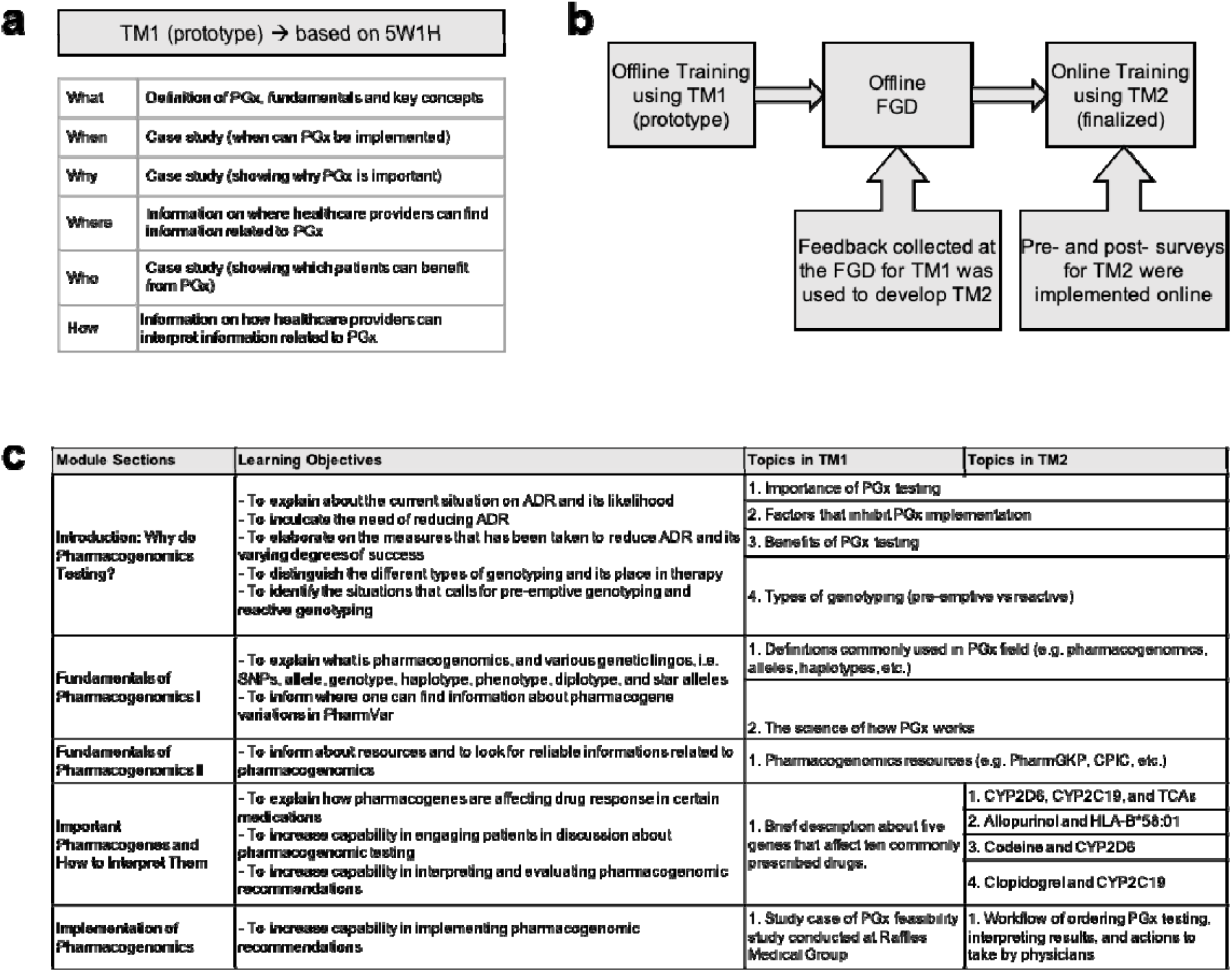
Development of training materials. **a**, Questions composed of 5W1H (what, when, why, where, who, and how) are used to determine the first training materials and objectives. **b**, The first training material (TM1) was designed based 5W1H questions as seen on Figure 1a and implemented in an offline training. Focus group discussion (FGD) with the TM1’s training participants was held offline to collect feedback. Feedback from the FGD was used to develop a more comprehensive training materials (TM2) and implemented as online training. **c**, Details of the learning objectives and topics covered in TM1 and TM2.

### Phase 1: Development of Training Materials

The content of both training materials focused on the PGx applications. Training outcomes were based on the competency inventory curated by the Pharmacogenetics/Pharmacogenomics Special Interest Group of the American Association of Colleges of Pharmacy [19]. The training objectives were: (1) to understand PGx applications to clinical practice, (2) to engage in patient discussion about PGx testing, and (3) to interpret, evaluate and implement PGx recommendations.

### Development of Training Material Prototype (TM1)

The “5W1H” approach was adopted for developing the initial training material. Elaborating on the “what, when, why, where, who and how” of PGx systematically introduced fundamental PGx concepts. The training began with “what” PGx is, defining fundamental terminology and key genetic concepts. Information on “where” HCPs may gather relevant PGx information and “how” to manoeuvre through PGx resources, such as PharmGKB, CPIC, and DPWG, was shared. A patient case was used to consolidate four concepts: “when” PGx testing can be implemented, “why” PGx is important, “how” HCPs can interpret PGx information, and “who” to apply PGx to in clinical settings.

### TM1 Evaluation: Focus Group Discussion (FGD)

TM1 was used to conduct a 30-minute duration offline training before the FGD. The main objective for the FGD was to collect feedback on TM1. The FGD spanned over an hour, was audio-recorded and conducted offline. FGD questions were tailored to explore the participants’ views on the fundamentals of PGx, PGx applications and course delivery. Questions were structured in an open-ended format to facilitate rich discussion. The FGD sequence of questions mirrored the sequence of TM1 content. Additionally, the free-flowing nature of the FGD allowed participants to voice suggestions on supplementary aspects of the training. Feedback collected during this FGD was used to revamp TM1 to TM2.

### TM2 Implementation

TM2 was used to conduct the final PGx implementation training. The final presentation utilizing TM2 was 90 minutes long due to changes in content and was done online via a private e-learning platform. This was due to the COVID-19 pandemic where social distancing was enforced. Therefore, online training was the most appropriate alternative. Pre-post surveys were distributed to participants before and after training respectively. The analysis of these surveys contributed to the validation of TM2.

### Phase 2: Training Evaluation Measures

We administered pre-training and post-training surveys to evaluate the training materials, both TM1 and TM2. Pre- and post-training surveys for TM1 were conducted offline while for TM2 were conducted online. FGD participants brought forward insightful comments regarding blockers to clinical pharmacogenomics implementation in the local context. Thus, feedback from the FGD was used to modify the survey questions in TM1 assessment and the modified survey questions were used to assess TM2, assessing the main blockers of clinical PGx implementation The surveys, gauging mostly parallel measures, were anonymous and unlinked. Written consent was obtained from the participants, highlighting voluntary participation. Survey questions were adapted from various studies analyzing HCPs’ general PGx perceptions [1,5] and HCPs’ attitudes and knowledge of existing PGx pre- and post-education [7,20]. Due to differing training objectives and varying content, specific modifications were made.

The survey consisted of five sections. Perceptions and self-efficacy sections consisted of two sub-sections (P1 and P2; SC1 and SC2) each and utilized five-point Likert scales. Knowledge questions required participants to choose the best multiple choice options. Needs assessment and evaluation of training sections were only included in the post-surveys. Rationales for each section are described below.

### Data Collection

Data collected was used to characterize the participants. To determine whether training could change clinical practice behaviour, the post-training survey also asked about experience with and anticipation of using PGx tests. An open ended section was incorporated in order to solicit feedback on the training course content and delivery to validate TM2 and facilitate future PGx educational programs.

Surveys incorporated the following aspects:

#### 1. Perceptions

To assess training objective (1), we evaluated for a perception change in clinical relevance (P1) and utility (P2) of PGx. Questions asked in this section are related to how useful PGx is towards the subjects’ clinical practice, and in what way PGx is useful.

#### 2. Self-efficacy

Evaluation of how to utilize PGx data in making drug therapy decisions (SE1) and how to engage in patient discussion about PGx (SE2) were necessary to assess training objectives (2) and (3). Questions asked in this section are related to how competent the subjects feel about implementing PGx practice.

#### 3. Knowledge

Knowledge, comprehension and application questions regarding clinical PGx recommendations were crafted as a patient case scenario to evaluate for training objective (3). Knowledge assessments were adapted from ASHP’s pharmacogenomics professional certification course. Questions assessing the knowledge taught in our developed PGx course were designed by licensed pharmacists who had undergone this ASHP’s certification course, this section also included a case study example. The aforementioned concepts were adopted from the first three levels of Bloom’s taxonomy, i.e. knowledge (remembering), comprehension (understanding), and application (applying).

### Statistical Analysis

Ordinal data related to participants’ perceptions and self-efficacy were summarized using median and interquartile range (IQR). Items assessed on a five-point Likert scale were collapsed and presented as the percentage of agree, disagree and neutral responses. Distribution of responses between the pre- and post-training surveys were compared using Mann-Whitney U test. Knowledge questions were scored as correct or incorrect, with missing items scored as incorrect. The percentage of correct responses overall and for each question on the pre- and post-training surveys were compared using chi-square test. Statistical analyses were conducted using R Version 3.5.2, with p < 0.05 considered as statistically significant.

## RESULTS

### Phase 1: Development of training materials

TM1 was implemented to train medical professionals who will be involved in a clinical study related to PGx implementation. Due to limited time given for offline training, more-detailed materials such as examples of drug-gene interactions were not explained in TM1. To develop TM2, feedback from training participants was collected through an offline focus group discussion (FGD). Feedback showed that the participants felt there were too few examples on common drug-gene interactions and on how PGx can be implemented in clinical settings. From the feedback received, TM2 was developed to have more in-depth materials, specifically for examples of drug-gene interactions commonly found in clinical settings. More details on the science of PGx and drug-gene interactions were also included in TM2. TM2 implementation was done online to have more flexibility in terms of time and place needed to complete the whole material, as well as scalability. (Fig 1)

### Characteristics of survey respondents from training

Overall, 102 respondents were collected in our study with 68 in TM1 and 34 in TM2. TM1 respondents consisted of 93.4% physicians, of which 68.9% mainly practiced in Family Medicine. On the other hand, TM2 consisted of two major groups of respondents, with majority being 61.8% physicians practicing Family Medicine and 33.3% pharmacy students. More than half of the respondents in TM1 (60.3%) and TM2 (70%) are experienced practitioners with more than 5 years of practice. Prior experience in PGx education is lacking across the respondents, only 27.9% in TM1 and 32.4% in TM2. This includes self-learning from independent resources (internet, colleague, journals, drug labels or package inserts), attending a lecture or seminar, and/or enrolling university curriculum. (Table 1)

**Table 1.**
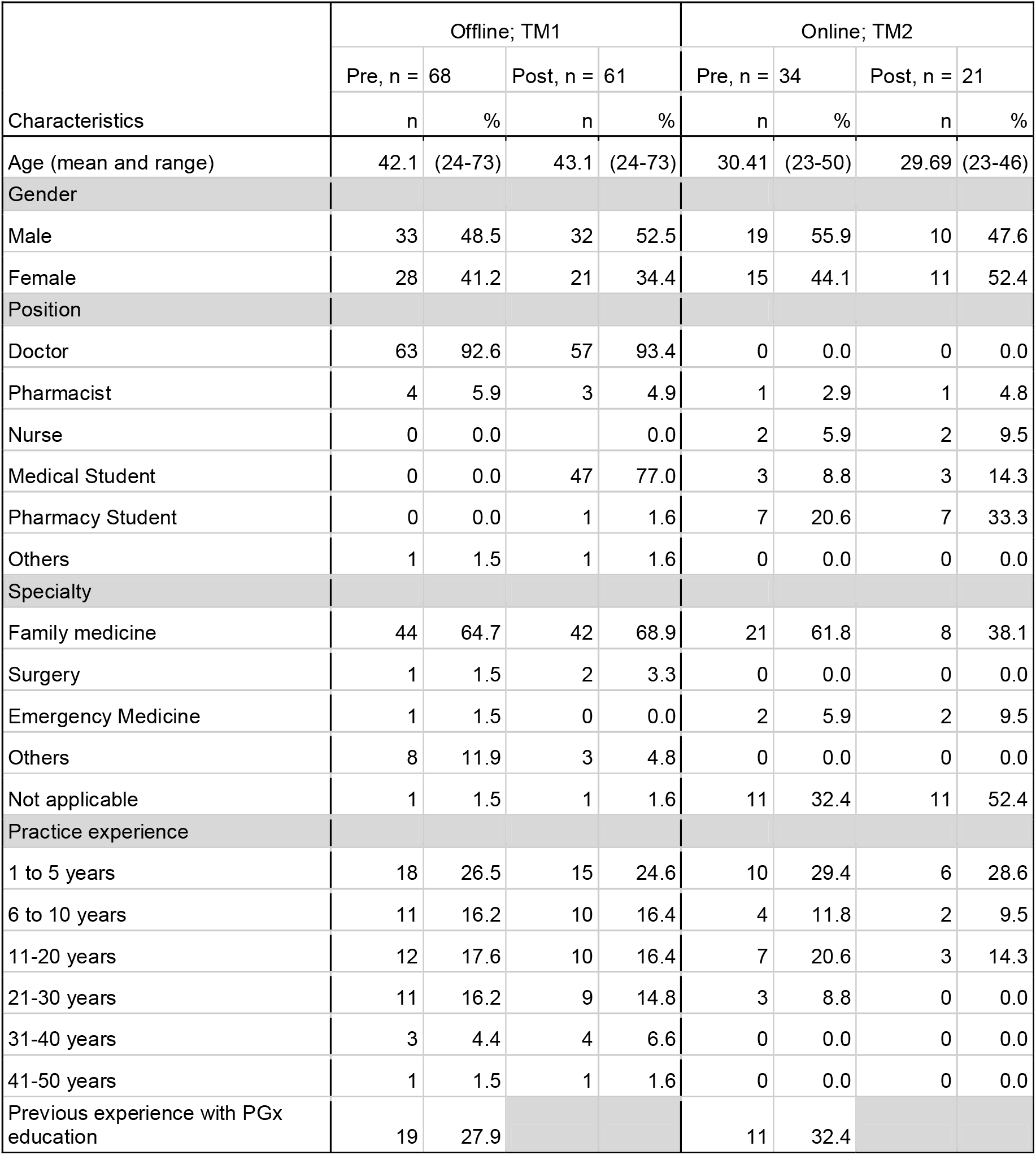
Participant Characteristics in Offline and Online Training.

### Relevance and utility of pharmacogenomics testing in clinical practice

To understand PGx applications in clinical practice, we evaluated for a perception change in clinical relevance and utility of PGx. This was done by inquiring a set of perception questions pre- and post-training, relative to 5-point Likert-type scale (Fig 2, Table 2). Prior to training, 52.4% in TM1 and 62.5% in TM2 participants generally agree or strongly agree on the clinical relevance and utility of PGx testing, indicating favorable perceptions towards PGx. This number increased even more after training to 84.8% in TM1 and 88.1% in TM2. Overall, participants’ median scoring in perceptions for TM1 improved from 3 to 4 (Mann-Whitney U test, p < 0.05), suggesting statistically significant positive perception change from TM1 training. While the corresponding median score for the online training remained at 4 from TM2 (Mann-Whitney U test, p < 0.05). Notably, 77% from TM1 and 85.7% from TM2 respondents reported greater anticipation of using PGx tests after attending the training.

**Table 2.**
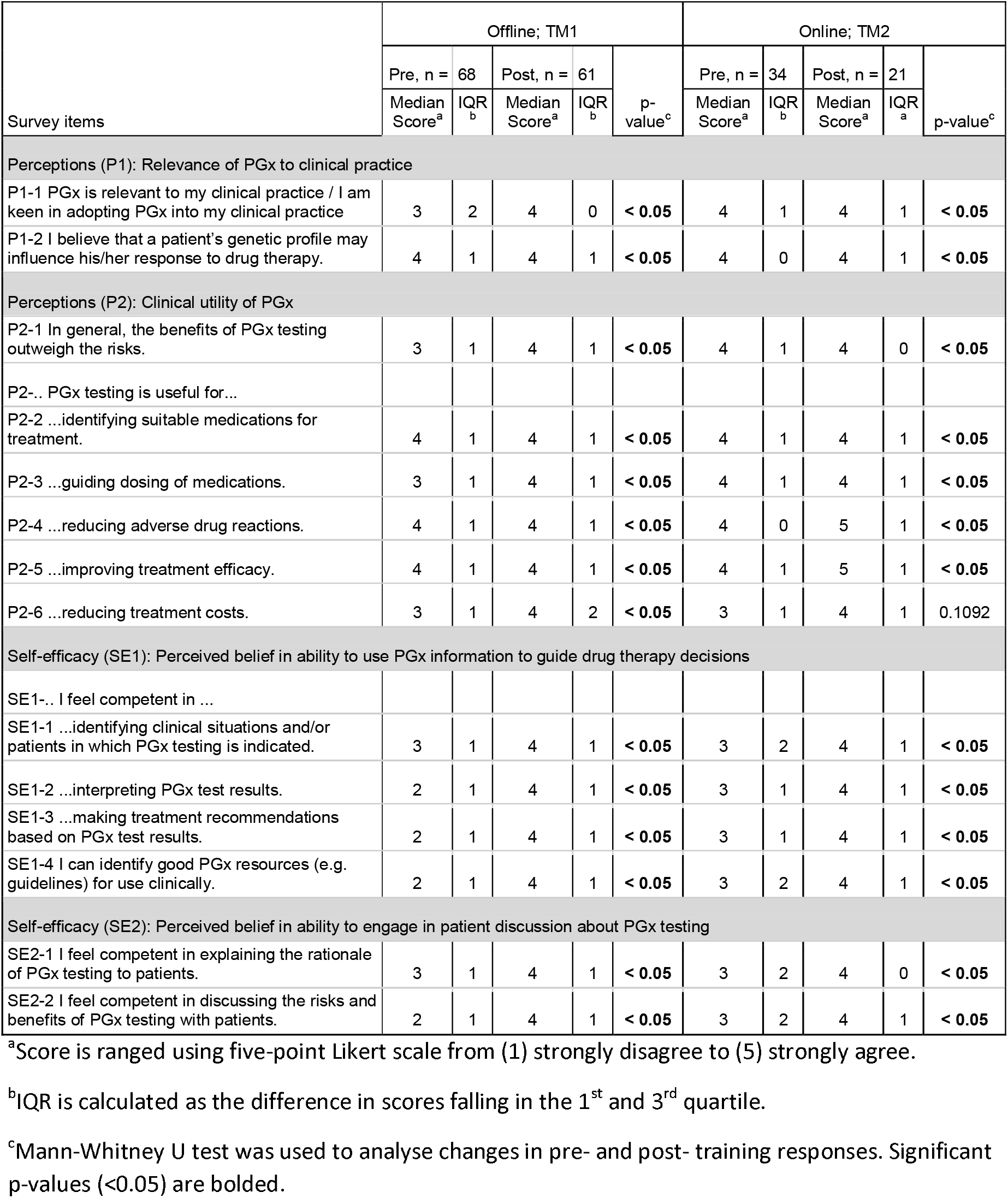
Pre- and post-training results related to perceptions and self-efficacy of addressing PGx testing.

**Fig. 2.**
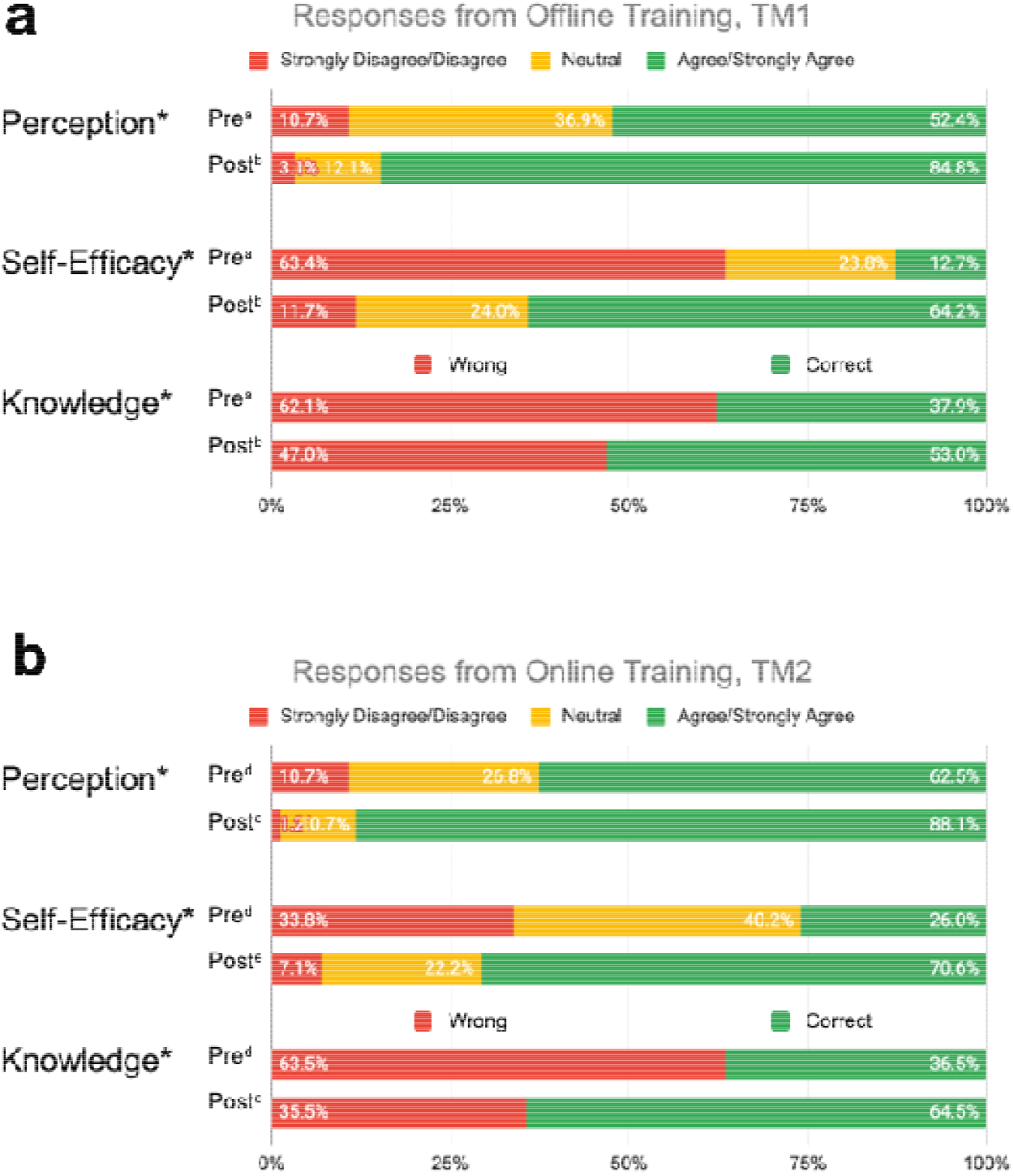
Overall percent of respondents relative to 5-point Likert-type scale labels pre- and post-PGx training conducted. **a**, Offline, TM1; and **b**, Online, TM2. Survey domains included Perceptions of the relevance and clinical utility of PGx, Self-Efficacy through perceived ability to use PGx information in guiding medical decisions and engage in patient discussions about PGx testing, and Knowledge questions on the PGx-based case studies. ^a^TM1 respondents pre-training, n = 68. ^b^TM1 respondents post-training, n = 61. ^c^TM2 respondents pre-training, n = 34. ^d^TM2 respondents post-training, n = 21. *Statistically significant p-values (p < 0.05) were observed in changes from pre- and post-training responses.

### Perceived ability in the implementation of pharmacogenomics in clinical setting

To evaluate the self-competency of respondents on the utilization of PGx data in making clinical decisions, we assess their ability to engage in PGx discussion, and consider PGx recommendations. We inquire on a set of self-efficacy questions on the perceived belief in ability to use PGx information to guide drug therapy decisions and engagement in patient discussion about PGx testing. Pre-training results demonstrate that respondents begin with inadequate self-efficacy in using PGx data to guide medication therapy and engage with patients (Fig 2, Table 2). Upon completion, both training increased their perceived ability in implementing PGx by 51.5% in TM1 and 44.6% in TM2. Participants median scoring in the self-efficacy section for both improved significantly from 2 to 4 in TM1, and 3 to 4 in TM2 (Mann-Whitney U test, p < 0.05).

### Knowledge and proficiency in applying pharmacogenomics to practice

Knowledge, comprehension and application questions were crafted as a patient case scenario to test the ability to interpret, evaluate and implement PGx recommendations following training. Questions were categorized under two sets – knowledge on theoretical PGx and practical clinical implementation of PGx. Respondents were quizzed pre- and post-training, and their performance was assessed to evaluate improvements (Fig 2, Table 3). On average, respondents significantly improved their correct response rate for proficiency questions by 15.1% in TM1 and 28.0% in TM2 (Chi-square test, p < 0.05). Both training have statistically significant improvements in scores for one knowledge level question under theoretical PGx category (Table 3). On the practical implementation of PGx, significant improvements are seen for an application level in TM1 and a comprehension level in TM2 (Table3).

**Table 3.** Correct responses to questions about knowledge on PGx comparing pre- and post-online training for TM1 and TM2.

| Survey Questions | Correct Answer | Correct Responses |  |  |  |  |  |  |  |  |  |
| --- | --- | --- | --- | --- | --- | --- | --- | --- | --- | --- | --- |
|  |  | Offline; TM1 |  |  |  |  | Online; TM2 |  |  |  |  |
|  |  | Pre |  | Post |  |  | Pre |  | Post |  |  |
|  |  | n | % | n | % |  | n | % | n | % |  |
| Knowledge (K1): Knowledge on theoretical PGx |  |  |  |  |  |  |  |  |  |  |  |
| K1-1. What may be the consequence of a PGx polymorphism? (comprehension level) | An individual has a higher risk for toxicity when using prescription drugs. | 40 | 59% | 43 | 65% | 0.4507<sup>a</sup> | 9 | 43% | 4 | 50% | 1<sup>b</sup> |
| K1-2. What does a poor metabolizer phenotype indicate? (knowledge level) | Decreased enzyme activity. | 17 | 25% | 18 | 27% | 0.7646<sup>a</sup> | 13 | 62% | 5 | 63% | 1<sup>b</sup> |
| K1-3. A patient with CYP2D6 activity score of 1.5 has which CYP2D6 phenotype? (knowledge level) | Normal metabolizer. | 6 | 38% | 10 | 63% | 0.1573<sup>a</sup> |  |  |  |  |  |
| K1-4. Which of the following is not correct about pre-emptive and reactive genotyping? (knowledge level) | Reactive genotyping has been shown to be more cost-effective than pre-emptive genotyping. | 14 | 27% | 28 | 56% | < 0.05<sup>a</sup> | 4 | 19% | 1 | 13% | 1<sup>b</sup> |
| K1-5. What does an ultra-rapid metabolizer phenotype for CYP2C19 indicate? (knowledge level) | Increased enzyme activity |  |  |  |  |  | 5 | 38% | 13 | 100% | < 0.05<sup>b</sup> |
| Knowledge (K2): Practical clinical implementation of PGx |  |  |  |  |  |  |  |  |  |  |  |
| K2-1. There is a high chance that Ms Lee will develop Stevens Johnsons Syndrome? (comprehension level) | FALSE |  |  |  |  |  | 8 | 62% | 12 | 92% | 0.1602<sup>b</sup> |
| K2-2. What is Ms Lee's CYP2D6 enzyme activity score? (comprehension level) | 2 |  |  |  |  |  | 2 | 15% | 13 | 100% | < 0.05<sup>a</sup> |
| K2-3. Which of the following would be appropriate regarding Ms Lee's amitriptyline therapy according to CPIC guidelines? (application level) | Consider alternative drug not metabolized by CYP2C19 |  |  |  |  |  | 1 | 8% | 1 | 8% | 1<sup>b</sup> |
| K2-4. A woman is diagnosed with breast cancer and, as part of her oncology regimen, she is treated with tamoxifen. She did not have genetic testing performed before initiating treatment. What PGx reason would cause the treating physician to decide to change the drug? (application level) | CYP2D6 poor metabolizer resulting in lack of drug. | 11 | 69% | 10 | 63% | 0.7097<sup>a</sup> |  |  |  |  |  |
| K2-5. Which of the following would be appropriate regarding clopidogrel therapy in patients who are CYP2C19 poor metabolizers? (application level) | Consider alternative antiplatelet therapy if no contraindications. | 15 | 29% | 31 | 62% | < 0.05<sup>a</sup> |  |  |  |  |  |

<sup>a</sup>Chi-square test was used to compare the percentage of correct responses between pre- and post-training surveys. Significant p-values (<0.05) are bolded.
<sup>b</sup>Fisher's test was used if criteria for Chi-square (expected value size > 5) is not met.

## DISCUSSION

This study evaluated the outcomes of PGx implementation training which was piloted at Continuing Education (CE) seminars and further developed into an online training module. The aim of the training was to educate respondents about the fundamental PGx theoretical concepts and clinical applications. Here, we found that respondents displayed positive perceptions of the clinical relevance and utility of PGx. We also demonstrated that PGx implementation training conducted as a case-based presentation can improve self-efficacy to clinically apply PGx information and to engage in PGx discussions.

There have been scarce resources available for doctors to learn pharmacogenomics online. The only accredited online pharmacogenomics certification course is offered by American Society of Health-System Pharmacists (ASHP). However, this course heavily focuses on how to set up a pharmacogenomics practice, i.e. sourcing labs to run samples and obtaining stakeholder approval. Our course aims to equip the enrollee with the most relevant clinical knowledge at the least amount of time, without going into the administrative details.

Prior to training, we observed that respondents portrayed positive perceptions of PGx. These results confirm the forward-thinking attitudinal findings gathered from other studies regarding clinicians’ belief in the concept of PGx and its potential benefits to improving drug efficacy and safety [5,14,17,21]. Another study also concluded that their hour-long PGx seminar significantly improved the attitudes of twelve physicians [7]. Advancements in PGx will continue and become more important for patient care to improve drug efficacy and safety [4,22]. Furthermore, their perceptions continued to improve significantly post-training, brightening prospects of increased PGx integration into clinical practice.

Conversely, pre-training responses demonstrate low self-efficacy amongst participants to integrate PGx into clinical practice. A knowledge translation gap of PGx use might explain their low perceived competency [5]. Despite efforts to increase PGx education in undergraduate and postgraduate healthcare courses [2,11–15,23,24], PGx educational efforts for clinicians are still not readily available [5,10,16]. Consequently, clinicians lack confidence in their knowledge of what tests are available, when and for whom to order, and how to interpret and incorporate PGx information to drug therapy [4,9]. Therefore, there is a need to advocate for PGx education available to HCP’s to improve their self-efficacy in practicing PGx.

Training significantly improved participants’ self-efficacy to integrate PGx into clinical practice. While clinical experience is associated with increased perceived competency [5], clinicians must already be proficient to provide PGx services when the opportunity arises [10,16]. Hence, it is important to improve clinicians’ self-efficacy to apply PGx to practice through education. Our finding indicates a continued need to improve PGx learning, to elevate clinicians’ perceived ability in using this therapeutic opportunity and encourage their prevalence in healthcare services.

There were improvements in knowledge questions when comparing pre- and post-training responses. Post-offline training, marginal improvements in participants’ knowledge were similarly observed after a PGx educational program for pharmacists [20]. On the other hand, post-online training significantly improved the correct response rate. Limited knowledge retention may have been the culprit in subpar improvements for the offline training due to the complexity of PGx or the transfer of overwhelming information over a short period [14,20]. Revisions made to TM2 addressed these pitfalls. Online training materials promote active learning [25] where participants can playback content to enhance knowledge retention. Moreover, complex PGx concepts were more thoroughly explained and quizzes helped to reinforce internalization of content. This is supported by our findings that all participants agreed that the quizzes helped in understanding PGx concepts. Additionally, the online training results reflect a more holistic improvement as the questions were formatted as a case scenario. This has been shown to be more effective for enhancing learning [25].

Similar to other studies [5,16,20], respondents express desire to learn more about PGx. Our study demonstrated 77% to 85.7% of respondents also anticipated using PGx tests after training. This highlights the need to develop effective PGx education for HCPs. We recommend introducing structured educational programs for HCPs to learn about PGx. Participants expressed eagerness to learn more about basic concepts of PGx and its application during the trainings. This highlights the need to develop sustainable education for HCPs. Furthermore, we suggest exploring hands-on development of PGx-focused clinical skills as it is vital to the effective adoption of PGx [5,8,14,20]. Hence, we recommend integrating point-of-care PGx information into electronic health records [5,26], and making PGx test kits readily available [5].

Most of the training materials that are available were developed in a top-down approach, without considering the audience’s level of knowledge. This study’s focus group discussion discovered that explaining pharmacogenetics concepts using case studies increases efficacy. Moreover, the delivery of training materials offline and online are increasingly important during the pandemic. This study shows that the combination of content, length, design, and platform to conduct training does not interfere with the efficacy of an offline training program. The study also highlights the importance of basic pharmacogenomics training for any healthcare professional interested in implementing it in their practice. Understanding the limitations of time and resources in new markets such as Asia, these training can be offered by teaching hospitals or commercial entities.

Our study has several possible limitations. In both training modules, the pre- and post-training survey responses were unlinked. Consequently, we could not analyze changes to individual responses and could only report aggregate data. Our study population were non-randomized and formed a convenient sample, which may imply selection bias for only respondents with PGx interests. While TM1 offline training involved mainly physicians, with limited participation from four pharmacists, TM2 online training had a lower response rate. This may limit the generalizability of our results to the broader population of clinicians. Results from pre- and post-surveys between TM1 and TM2 could also have been impacted by the mode of delivery (offline vs online). Finally, actual implementation of testing and the long-term effects of training were not evaluated. This is because our study was intended to provide baseline and initial assessments of the outcomes of PGx implementation training for clinicians. Therefore, we suggest conducting future studies to follow HCP’s over a prolonged period to evaluate the effectiveness of regular PGx educational programs and actual clinical update of PGx integration. A follow-up study is currently underway with our webinar respondents to assess the sustainability of the training’s impacts during the clinical implementation of PGx in their practice.

## CONCLUSION

Overall, respondents have favorable perceptions towards PGx testing, but lack self-efficacy and competency in PGx data utilization. Training has been proven to significantly improve self-efficacy and competency, and lead to greater anticipation of PGx adoption in clinical practice. Online training delivery mode is evidently preferable for further improvements. With its flexibility and scalability, it can be expanded as continuous education over a prolonged period to evaluate the effectiveness of PGx education and integration into clinical practice.

## FUTURE PERSPECTIVE

Online training delivery mode is evidently preferable for further improvements. With its ability to be flexible and scalable, it can be expanded as continuous education over a prolonged period to evaluate the effectiveness of PGx education and integration into clinical practice.

## EXECUTIVE SUMMARY

### Background

- PGx testing has demonstrated potential to enhance prevention of ADR occurrence in patients with distinguishable genetic variations.
- However, due to lack of PGx education among HCPs, testing is not administered regularly in clinical practice.
- PGx education has only seen increase in medical schools, but not in practicing clinicians.

### Methods

- Training materials were developed to cover theoretical concepts and clinical applications of PGx as implementation training.
- PGx implementation training was conducted across diverse group of HCPs including clinicians, pharmacists and students from both fields.
- Evaluate changes in thoughts and opinions of respondents pre- and post-training.

### Results

- Pre-training responses demonstrate respondents already have positive perception of the utility of PGx testing.
- Respondents have statistically significant increase in self-assessed efficacy and PGx knowledge upon completion of training.

### Conclusion

- PGx education improve HCPs self-assessment and encourage adoption of PGx diagnostics to support clinical decisions.
- Online training delivery mode is preferred for their flexibility and scalability as continuous education.

## Data Availability

Will individual participant data be available (including data dictionaries)?
Yes
What data in particular will be shared?
Individual participant data that underlie the results reported in this article, after deidentification (text, tables, figures, and appendices).
What other documents will be available?
None
When will data be available (start and end dates)?
Immediately following publication. No end date.
With whom?
Anyone who wishes to access the data.
For what types of analyses?
Any purpose.
By what mechanism will data be made available?
All data generate or analysed during this study are included in this published article and appendices. Request for additional material should be addressed to A.I.

## AUTHOR CONTRIBUTIONS

F.A. and A.I. conceived the study. S.G. and A.M.S.R. designed survey and developed training material with guidance from F.A., L.S., A.C. and A.I. F.A., A.C. and A.I. conducted the trainings. C.M. analysed data collected. C.M. and K.I.J wrote the manuscript with input from all authors

## ACKNOWLEDGEMENTS

We thank the participants for their time in attending the training courses, focus group discussions and filling our survey. We thank S. Chandrasekaran and M. Tan for assistance in developing the training materials.

## ETHICAL DISCLOSURE

This study was approved by the institutional review board (IRB No. 038/KEPK/III/2018 for Indonesia; 2017/007 for Singapore). Written consent was obtained from FGD participants, highlighting voluntary participation.

## DATA SHARING STATEMENT

**Will individual participant data be available (including data dictionaries)?**

Yes

**What data in particular will be shared?**

Individual participant data that underlie the results reported in this article, after deidentification (text, tables, figures, and appendices).

**What other documents will be available?**

None

**When will data be available (start and end dates)?**

Immediately following publication. No end date.

**With whom?**

Anyone who wishes to access the data.

**For what types of analyses?**

Any purpose.

**By what mechanism will data be made available?**

All data generate or analysed during this study are included in this published article and appendices. Request for additional material should be addressed to A.I.

## Appendix 1

Pre-training survey for TM1

### Survey 1: Pre-training pharmacogenomics survey

#### Introduction

Pharmacogenomics is the study of how genes affect an individual’s response to drugs. This survey aims to examine clinicians’ knowledge and perceptions of the clinical use of pharmacogenomics testing. This survey should take approximately 5 minutes to complete; thank you for your time.

Please rate the following (circle your answer):

#### Section 1: Perceptions

1. Pharmacogenomics is useful to my current practice.

| Strongly Disagree |  |  | Strongly Agree |  |
| --- | --- | --- | --- | --- |
| 1 | 2 | 3 | 4 | 5 |

2. I believe that a patient’s genetic profile may influence his/her response to drug therapy.

2. I believe that a patient's genetic profile may influence his/her response to drug therapy.
| Strongly Disagree |  |  | Strongly Agree |  |
| --- | --- | --- | --- | --- |
| 1 | 2 | 3 | 4 | 5 |

3. In general, the benefits of pharmacogenomics testing outweigh the risks.

| Strongly Disagree |  |  | Strongly Agree |  |
| --- | --- | --- | --- | --- |
| 1 | 2 | 3 | 4 | 5 |

4. Is pharmacogenomics testing useful for the following?

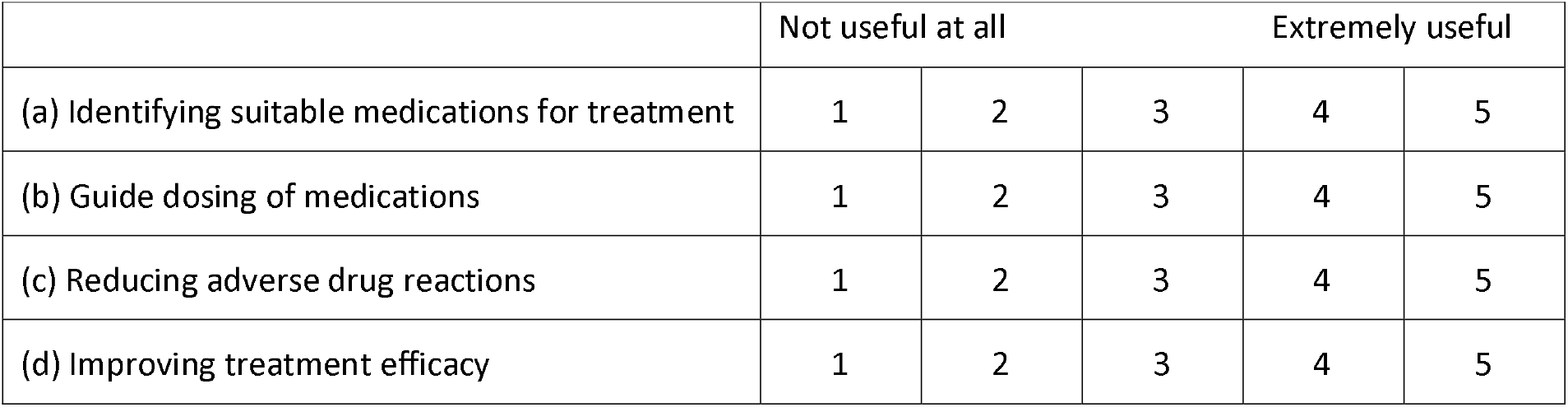
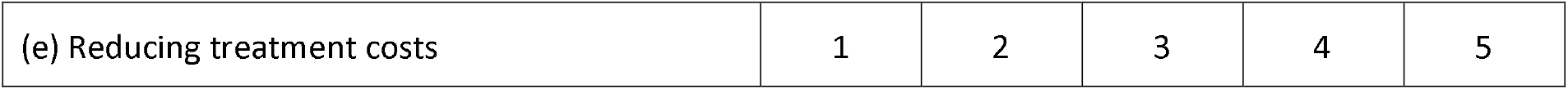

|  | Not useful at all |  |  | Extremely useful |  |
| --- | --- | --- | --- | --- | --- |
| (a) Identifying suitable medications for treatment | 1 | 2 | 3 | 4 | 5 |
| (b) Guide dosing of medications | 1 | 2 | 3 | 4 | 5 |
| (c) Reducing adverse drug reactions | 1 | 2 | 3 | 4 | 5 |
| (d) Improving treatment efficacy | 1 | 2 | 3 | 4 | 5 |
| (e) Reducing treatment costs | 1 | 2 | 3 | 4 | 5 |

#### Section 2: Ability

5. Please rank your perceived ability:

a. I feel competent in identifying clinical situations and/or patients in which pharmacogenomics testing is indicated.

b. I feel competent in interpreting results of pharmacogenomics tests.

|  |  |  |  |  |
| --- | --- | --- | --- | --- |
| Strongly Disagree |  |  | Strongly Agree |  |
| 1 | 2 | 3 | 4 | 5 |

c. I feel competent in making treatment recommendations based on results.

(b) I feel competent in interpreting results of pharmacogenomics tests.
|  |  |  |  |  |
| --- | --- | --- | --- | --- |
| Strongly Disagree |  |  | Strongly Agree |  |
| 1 | 2 | 3 | 4 | 5 |

d. I can identify good pharmacogenomics resources (e.g. guidelines) for use clinically.

(c) I feel competent in making treatment recommendations based on results.
|  |  |  |  |  |
| --- | --- | --- | --- | --- |
| Strongly Disagree |  |  | Strongly Agree |  |
| 1 | 2 | 3 | 4 | 5 |

e. I feel competent in explaining the rationale of pharmacogenomics testing to patients.

(d) I can identify good pharmacogenomics resources (e.g. guidelines) for use clinically.
|  |  |  |  |  |
| --- | --- | --- | --- | --- |
| Strongly Disagree |  |  | Strongly Agree |  |
| 1 | 2 | 3 | 4 | 5 |

f. I feel competent in discussing the risks and benefits of pharmacogenomics testing with patients.

(e) I feel competent in explaining the rationale of pharmacogenomics testing to patients.
|  |  |  |  |  |
| --- | --- | --- | --- | --- |
| Strongly Disagree |  |  | Strongly Agree |  |
| 1 | 2 | 3 | 4 | 5 |

(f) I feel competent in discussing the risks and benefits of pharmacogenomics testing with patients.
|  |  |  |  |  |
| --- | --- | --- | --- | --- |
| Strongly Disagree |  |  | Strongly Agree |  |
| 1 | 2 | 3 | 4 | 5 |

#### Section 3: Knowledge

6. What may be the consequence of a pharmacogenomics polymorphism?

- An individual cannot metabolize any drugs
- An individual has a higher risk for toxicity when using prescription drugs
- A single drug dose is appropriate for a given indication
- Individualized dose adjustments should be made according to body surface area

7. What does a poor metabolizer phenotype indicate?

- Lower drug safety because of poor metabolism
- Good drug efficacy because of poor metabolism
- Decreased enzyme activity
- Increased enzyme activity

8. Which of the following is **<u>no</u>t** correct about pre-emptive and reactive genotyping?

- Reactive genotyping is ordered as a drug therapy is being initiated or contemplated.
- Pre-emptive genotyping allows pharmacogenomics information to be available to guide prescribing.
- Reactive genotyping has been shown to be more cost-effective than pre-emptive genotyping.
- Pre-emptive genotyping usually tests for a panel of genes, whereas reactive genotyping usually tests for one to two genes.

9. Which of the following would be appropriate regarding clopidogrel therapy in patients who are CYP2C19 poor metabolizers?

- Initiate therapy with recommended starting dose
- Consider a 25% increase of recommended starting dose
- Consider a 25% decrease of recommended starting dose
- Consider alternative antiplatelet therapy if no contraindications

10. What sources have you used to learn about pharmacogenomics testing and its applications? (please select all that apply)

- Undergraduate education curriculum
- Postgraduate education curriculum
- Internet, website:__________
- Seminar, seminar name:_____________
- Journal, journal name:________________
- Drug labels (package inserts)
- Colleague
- I have not learnt about pharmacogenomics testing and its applications
- Other (please specify):_____________

#### Section 4: Demographics

11. Year of birth:

12. Gender: Male Female

13. Position:

- Doctor
- Pharmacist
- Nurse
- Other (please specify):____________

14. Main practice specialty:_____________

15. Number of years of practice experience:_____________________

## Appendix 2 Post-training survey for TM1

### Survey 2: Post-training pharmacogenomics survey

Please rate the following (circle your answer):

#### Section 1: Perceptions

1 Pharmacogenomics is useful to my current practice.

| Strongly Disagree |  |  | Strongly Agree |  |
| --- | --- | --- | --- | --- |
| 1 | 2 | 3 | 4 | 5 |

2. I believe that a patient’s genetic profile may influence his/her response to drug therapy.

2. I believe that a patient's genetic profile may influence his/her response to drug therapy.
| Strongly Disagree |  |  | Strongly Agree |  |
| --- | --- | --- | --- | --- |
| 1 | 2 | 3 | 4 | 5 |

3. In general, the benefits of pharmacogenomics testing outweigh the risks.

| Strongly Disagree |  |  | Strongly Agree |  |
| --- | --- | --- | --- | --- |
| 1 | 2 | 3 | 4 | 5 |

4. Is pharmacogenomics testing useful for the following?

|  | Not useful at all |  |  | Extremely useful |  |
| --- | --- | --- | --- | --- | --- |
| (a) Identifying suitable medications for treatment | 1 | 2 | 3 | 4 | 5 |
| (b) Guide dosing of medications | 1 | 2 | 3 | 4 | 5 |
| (c) Reducing adverse drug reactions | 1 | 2 | 3 | 4 | 5 |
| (d) Improving treatment efficacy | 1 | 2 | 3 | 4 | 5 |
| (e) Reducing treatment costs | 1 | 2 | 3 | 4 | 5 |

#### Section 2: Ability

Please rank your perceived ability:

b. I feel competent in interpreting results of pharmacogenomics tests.

| Strongly Disagree |  |  | Strongly Agree |  |
| --- | --- | --- | --- | --- |
| 1 | 2 | 3 | 4 | 5 |

c. I feel competent in making treatment recommendations based on results.

(b) I feel competent in interpreting results of pharmacogenomics tests.
| Strongly Disagree |  |  | Strongly Agree |  |
| --- | --- | --- | --- | --- |
| 1 | 2 | 3 | 4 | 5 |

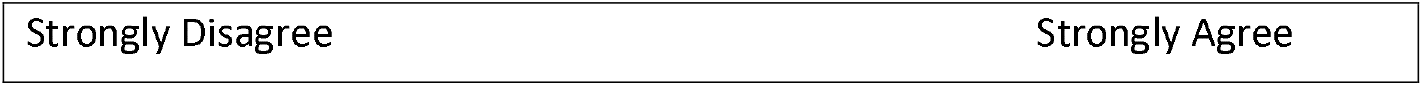
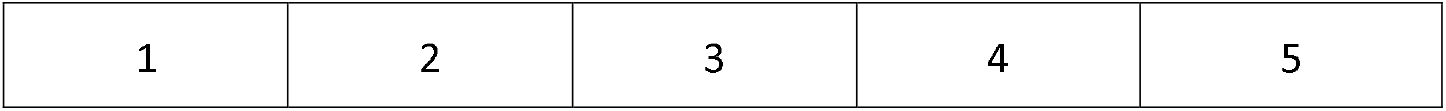
d. I can identify good pharmacogenomics resources (e.g. guidelines) for use clinically.

(c) I feel competent in making treatment recommendations based on results.
| Strongly Disagree |  |  | Strongly Agree |
| --- | --- | --- | --- |

|  |  |  |  |  |
|---|---|---|---|---|
| 1 | 2 | 3 | 4 | 5 |

e. I feel competent in explaining the rationale of pharmacogenomics testing to patients.

(d) I can identify good pharmacogenomics resources (e.g. guidelines) for use clinically.
|  |  |  |  |  |
| --- | --- | --- | --- | --- |
| Strongly Disagree |  |  | Strongly Agree |  |
| 1 | 2 | 3 | 4 | 5 |

f. I feel competent in discussing the risks and benefits of pharmacogenomics testing with patients.

(e) I feel competent in explaining the rationale of pharmacogenomics testing to patients.
|  |  |  |  |  |
| --- | --- | --- | --- | --- |
| Strongly Disagree |  |  | Strongly Agree |  |
| 1 | 2 | 3 | 4 | 5 |

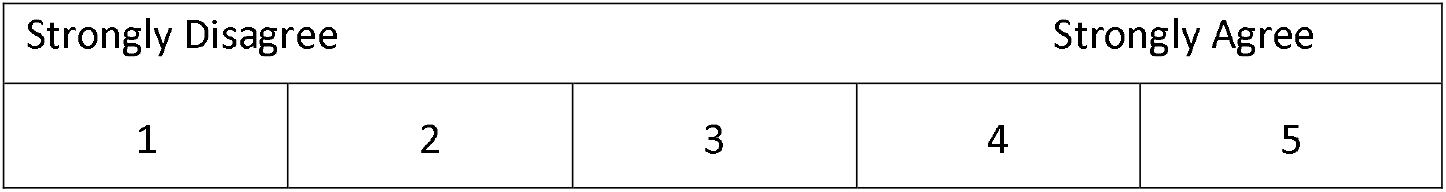

(f) I feel competent in discussing the risks and benefits of pharmacogenomics testing with patients.
|  |  |  |  |  |
| --- | --- | --- | --- | --- |
| Strongly Disagree |  |  | Strongly Agree |  |
| 1 | 2 | 3 | 4 | 5 |

#### Section 3: Knowledge

5. What may be the consequence of a pharmacogenomics polymorphism?

6. What does a poor metabolizer phenotype indicate?

7. Which of the following is **<u>not</u>** correct about pre-emptive and reactive genotyping?

8. Which of the following would be appropriate regarding clopidogrel therapy in patients who are CYP2C19 poor metabolizers?

#### Section 4: Needs assessment

9. To better utilize pharmacogenomics information in the management of drug therapy, I would need… (please select all that apply)

- Better knowledge on pharmacology
- Better knowledge on drug metabolism
- Better knowledge on the basic concepts of pharmacogenomics
- Stronger evidence that pharmacogenomics improves clinical outcomes
- Better ability to apply my knowledge
- Better knowledge of legal regulations
- Support of my working institution
- Insurance coverage
- Expert counsel
- Other (please specify):_____________

10. What is your preferred format for learning more about pharmacogenomics? (please select all that apply)

- Lectures
- Journal clubs
- Medical app
- E-learning
- Case discussion
- Other (please specify):_________________

#### Section 5: Evaluation of the training

11. Please rate the following:

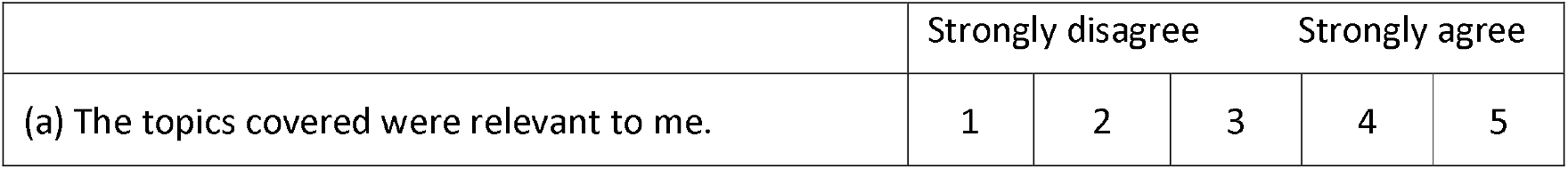
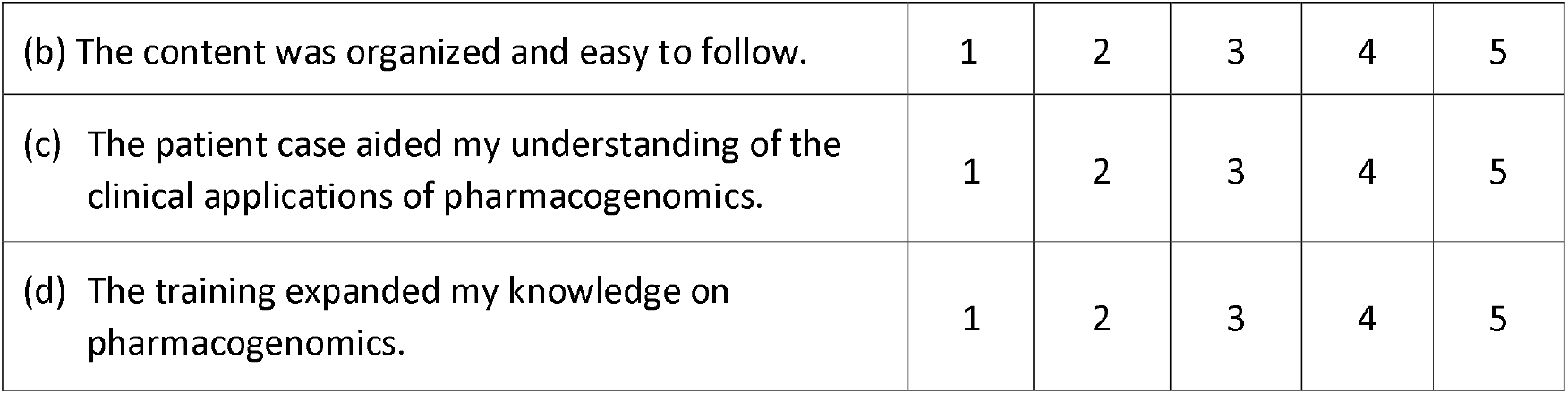

#### Section 6: Demographics

12. Within the past 6 months, how often have you ordered or recommended a pharmacogenomics test?

- 0
- 1 time per month
- 2-5 times per month
- >5 times per month

13. Do you anticipate ordering or recommending a pharmacogenomics test for a patient within the next 6 months?

- Yes
- No

14. Year of birth:________

15. Gender: Male Female

16. Position:

- Doctor
- Pharmacist
- Nurse
- Other (please specify):_____________

17. Main practice specialty:

18. Number of years of practice experience:

19. Please list any additional comments or feedback here:

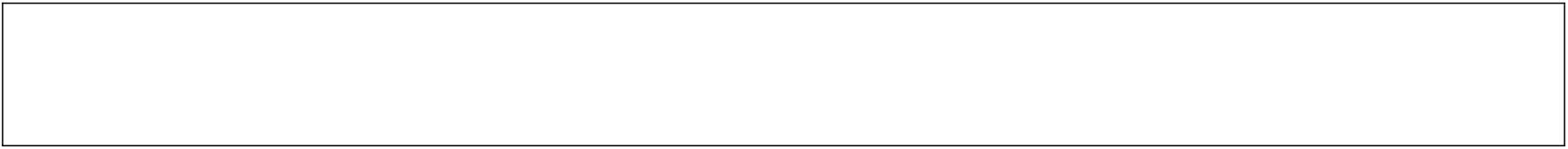

## Appendix 3

Pre-training survey for TM2

### Nalagenetics Pharmacogenomics Pre-Training Survey

We are Nalagenetics and we are creating a pharmacogenomics (PGx) program to integrate PGx into clinical practice. Pharmacogenomics is the study of how genes affect an individual’s response to drugs. One of the main benefits of PGx is to prevent and minimize adverse drug reactions (ADRs). This survey aims to gain feedback on our existing prototype PGx course. It will take approximately 10 minutes to complete, thank you for your time.

*Required

#### Perceptions

1. Are you currently integrating PGx into your clinical practice? *

a. Yes
b. No

2. Please rate the following: * (Mark only one oval per row.)

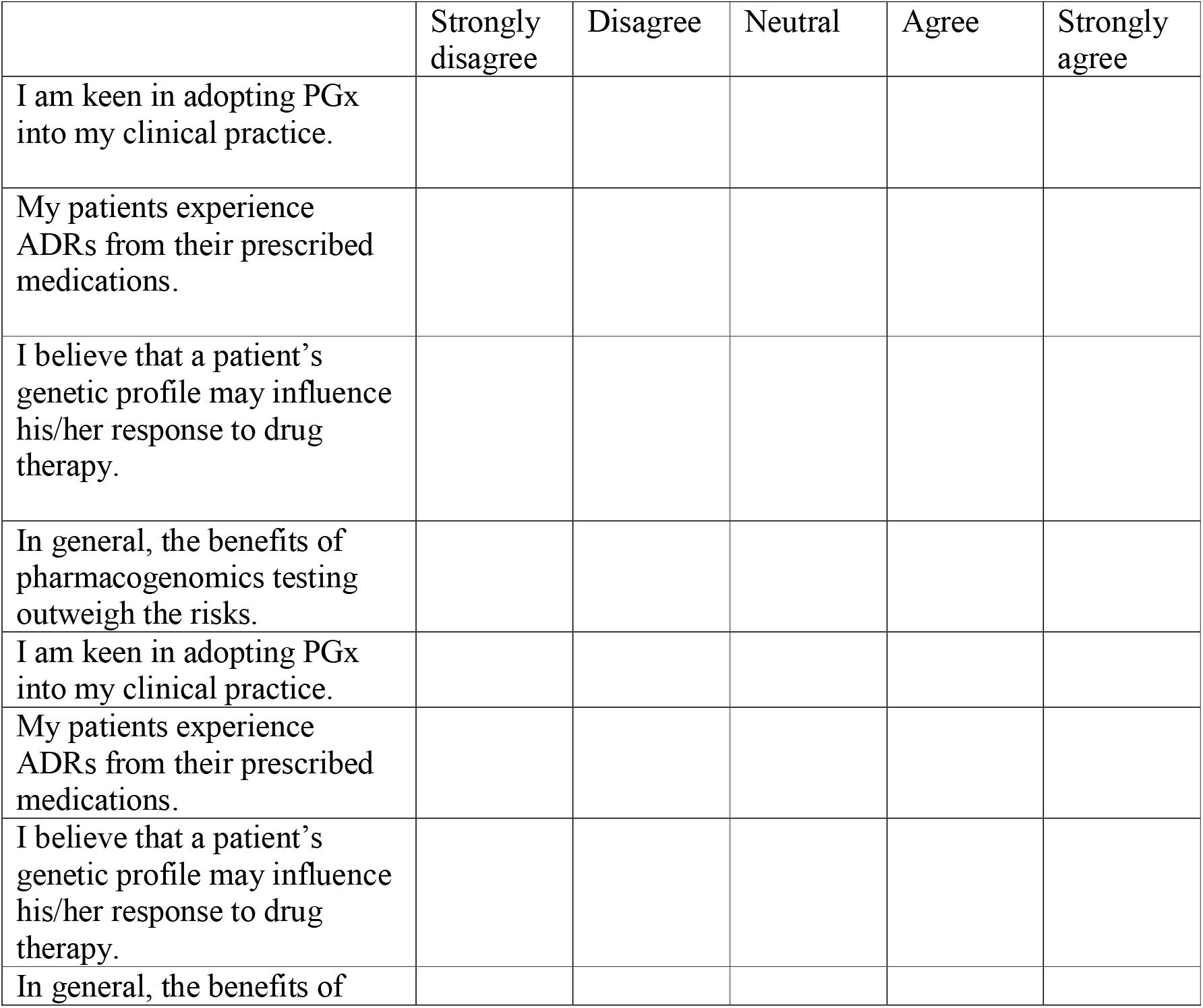
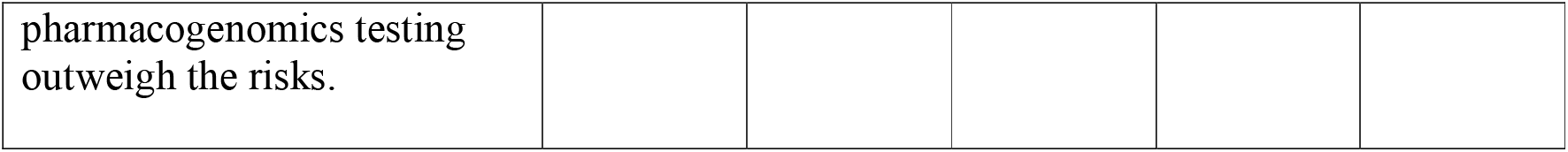

3. PGx is useful for the following: * (Mark only one oval per row.)

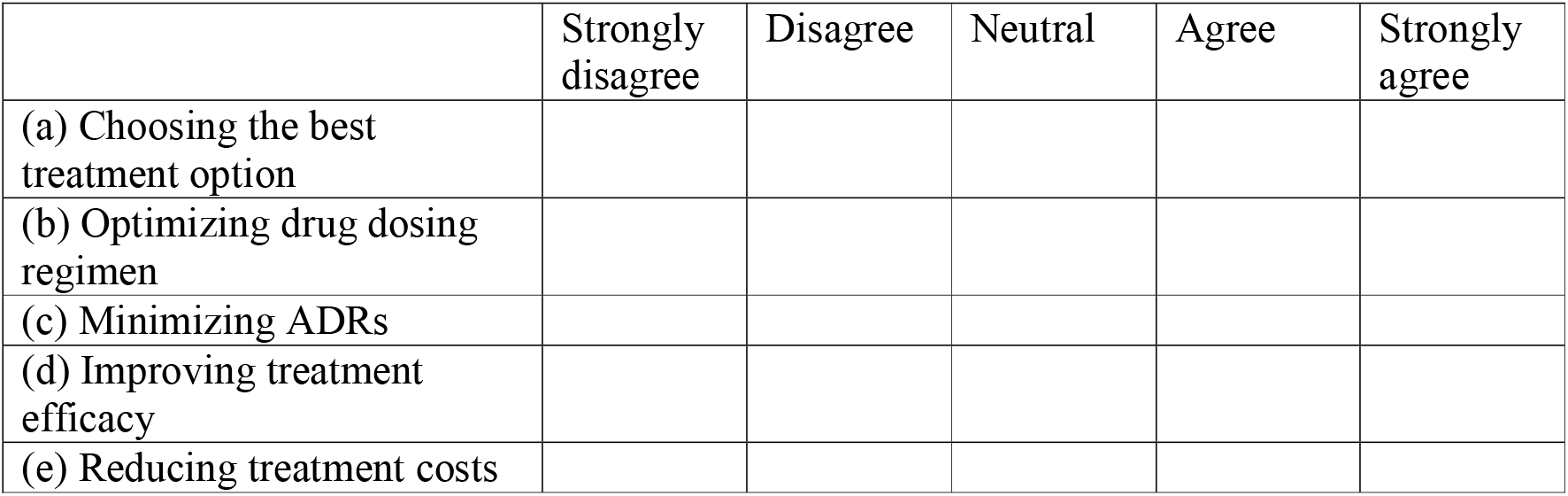

4. Some of the obstacles involved in implementing PGx into my clinical practice are: * (Mark only one oval per row.)

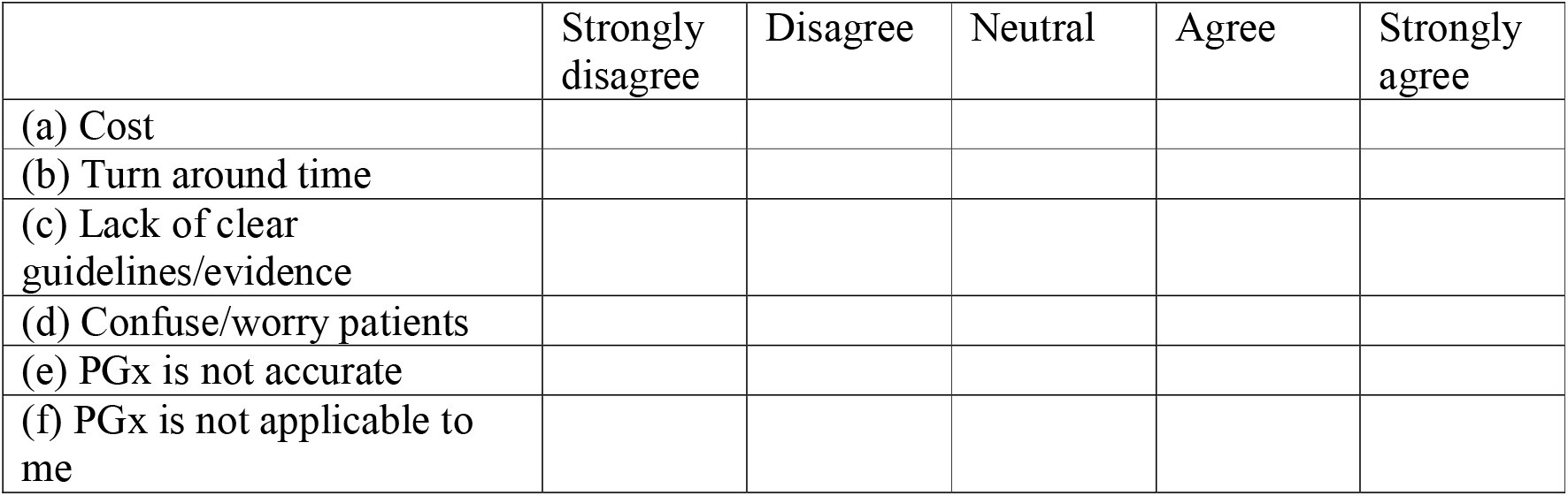

#### Ability

5. If I were to adopt PGx into my clinical practice, I know how to do the following: * (Mark only one oval per row.)

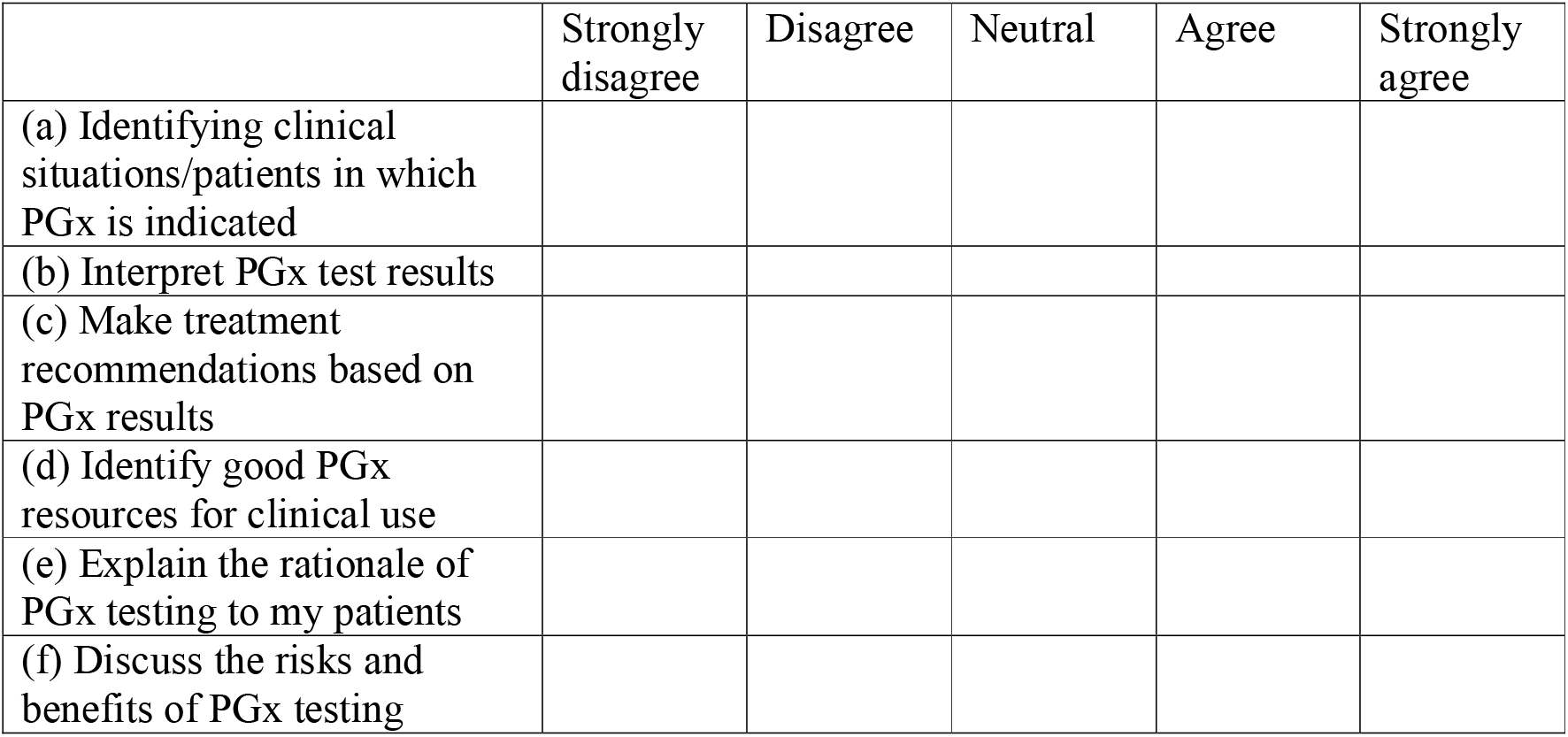

#### Knowledge

6. What may be the consequence of a pharmacogenomics polymorphism? (Mark only one oval.)

a. An individual cannot metabolize any drugs
b. An individual has a higher risk for toxicity when using prescription drugs
c. A single drug dose is appropriate for a given indication
d. Individualized dose adjustments should be made according to body surface area

7. What does a poor metabolizer phenotype indicate? (Mark only one oval.)

a. Lower drug safety because of poor metabolism
b. Good drug efficacy because of poor metabolism
c. Decreased enzyme activity
d. Increased enzyme activity

8. Which of the following is not correct about pre-emptive and reactive genotyping? (Mark only one oval.)

a. Reactive genotyping is ordered as a drug therapy is being initiated or contemplated.
b. Pre-emptive genotyping allows pharmacogenomics information to be available to guide prescribing.
c. Reactive genotyping has been shown to be more cost-effective than pre-emptive genotyping.
d. Pre-emptive genotyping usually tests for a panel of genes, whereas reactive genotyping usually tests for one to two genes.

#### Evaluation

9. Please rate the following: * (Mark only one oval per row.)

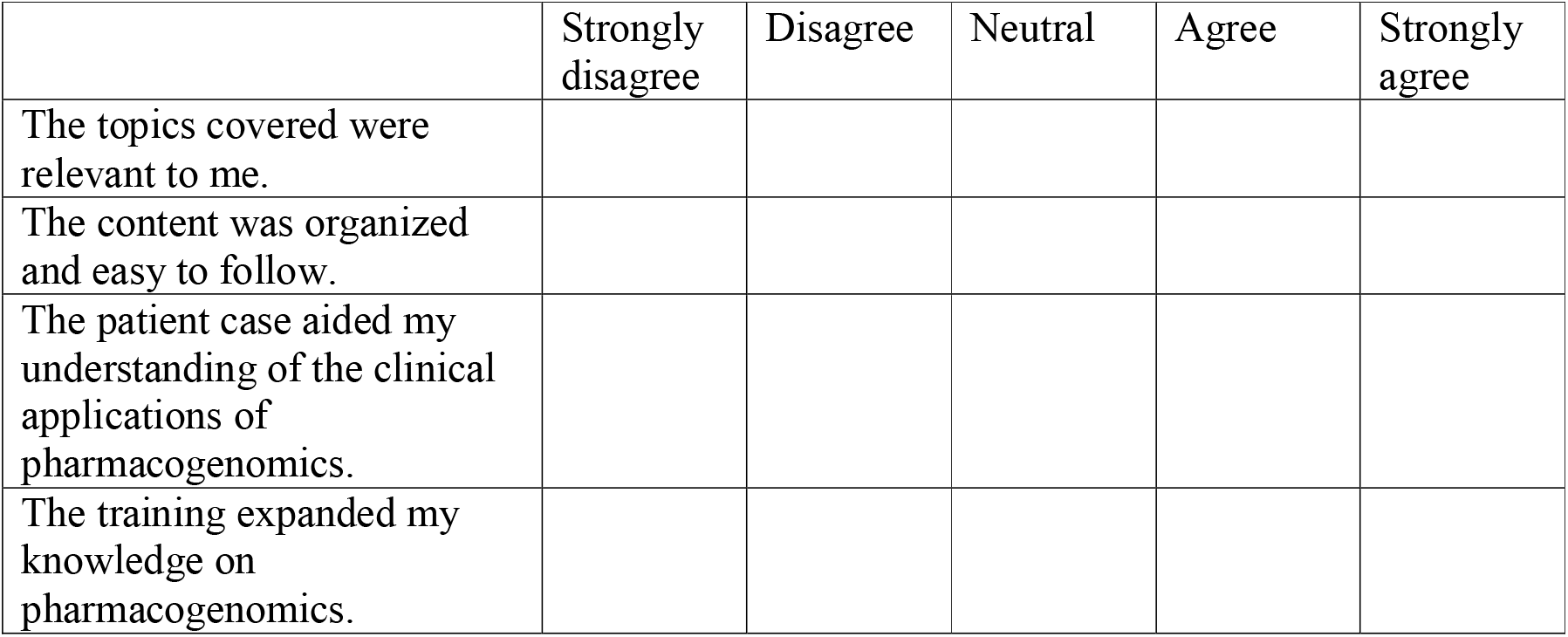

## Appendix 4

Post-training survey for TM2

### Nalagenetics Pharmacogenomics Post-Training Survey

*Required

1. Age Group: * Mark only one oval.

a. <30
b. 30-39
c. c. 40-49
d. d. 50-59
e. e. >60

2. Gender * Mark only one oval.

a. Female
b. Male
c. Prefer not to say
d. Other:

3. Number of practicing years * Mark only one oval.

a. <5
b. 5-10
c. 11-20
d. 21-30
e. >30

4. Where did you study medicine? * Mark only one oval.

a. Singapore
b. Overseas

5. What is your specialty?

6. I learnt about pharmacogenomics during my medical school * Mark only one oval.

a. Yes
b. No
c. I am not sure

#### Perceptions

7. Are you currently integrating PGx into your clinical practice? * Mark only one oval.

a. Yes
b. No

8. Please rate the following: * Mark only one oval per row.

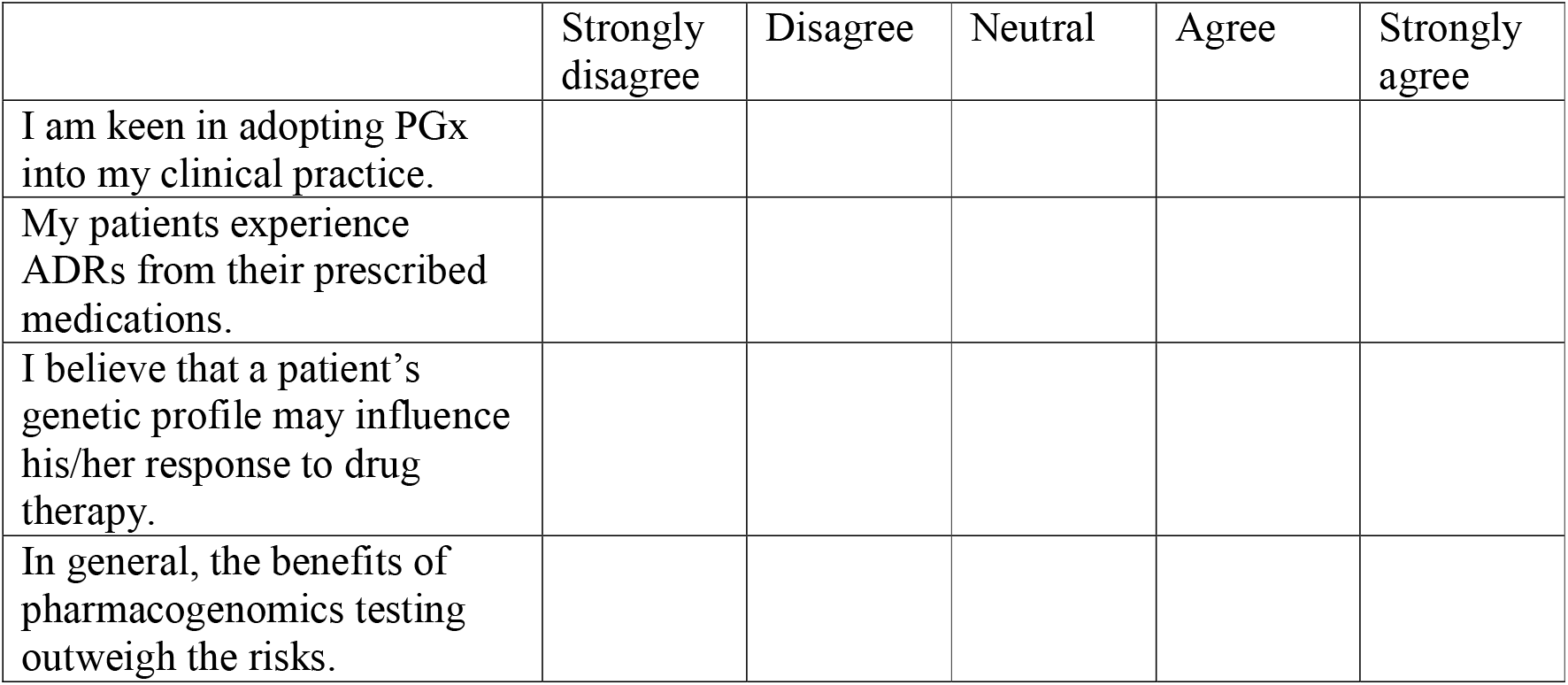

9. PGx is useful for the following: * Mark only one oval per row.

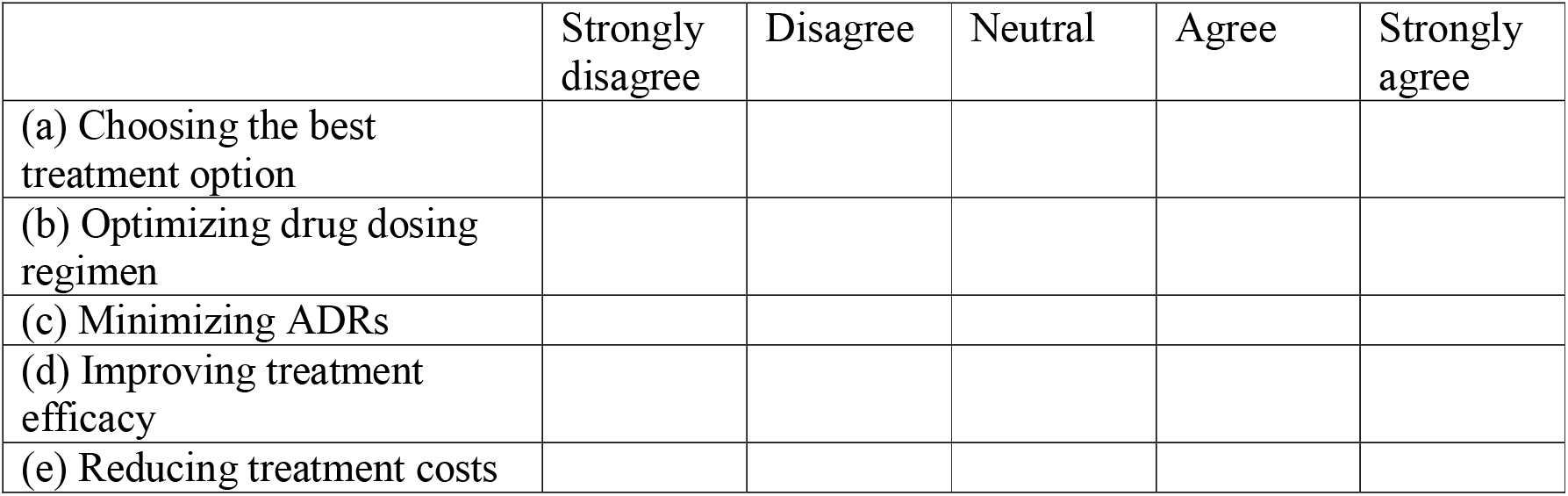

10. Some of the obstacles involved in implementing PGx into my clinical practice are: * Mark only one oval per row.

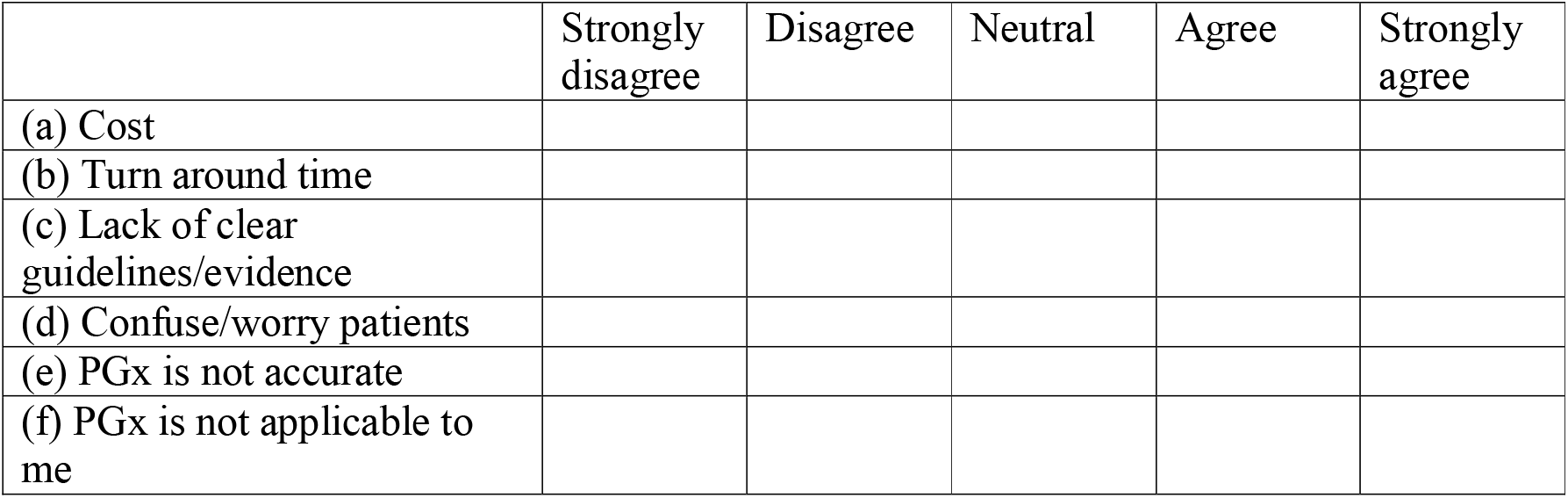

#### Ability

11. If I were to adopt PGx into my clinical practice, I know how to do the following: * Mark only one oval per row.

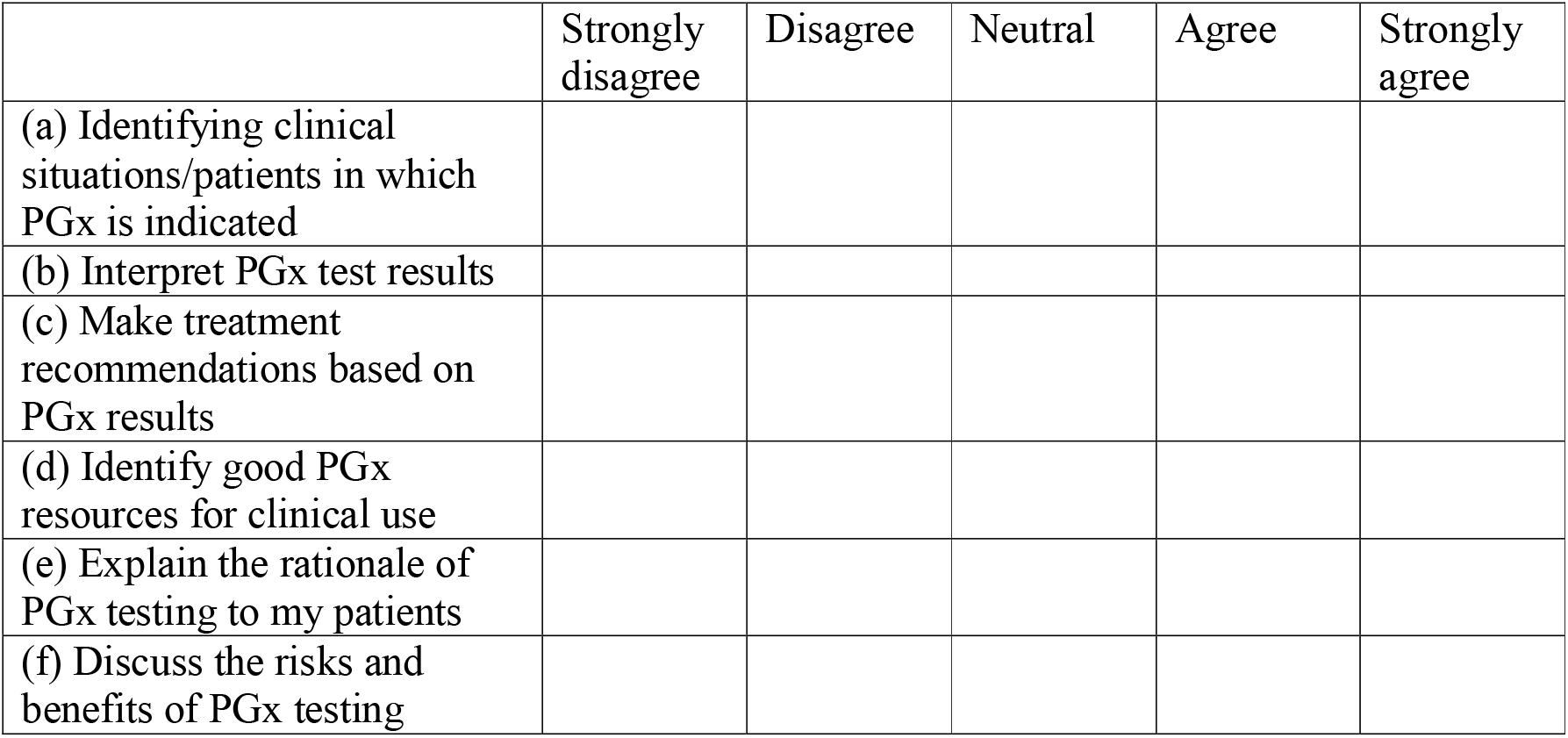

#### Knowledge

12. What may be the consequence of a pharmacogenomics polymorphism? Mark only one oval.

13. What does a poor metabolizer phenotype indicate? Mark only one oval.

14. Which of the following is not correct about pre-emptive and reactive genotyping? Mark only one oval.

#### Evaluation

15. Please rate the following: * Mark only one oval per row.

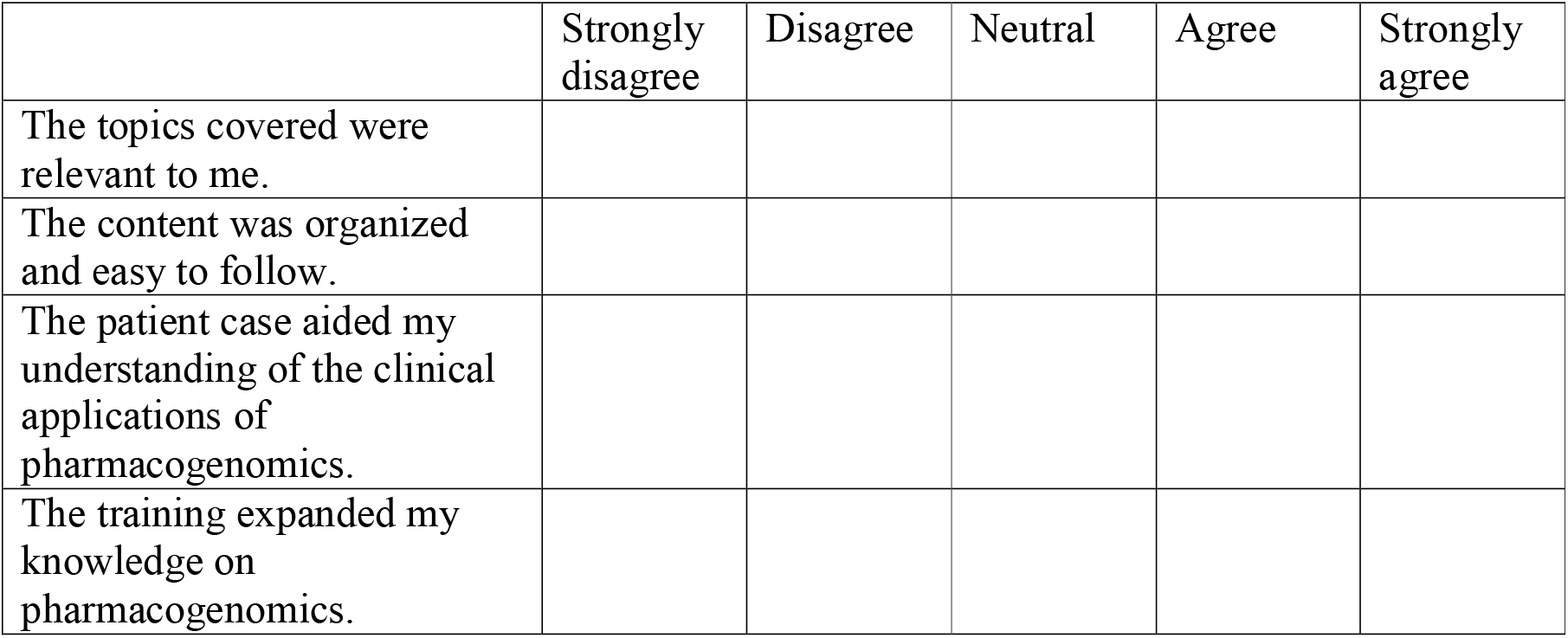

## Appendix 5

TM2 Course content

**Online course can be found here:** https://learning.nalagenetics.com/courses/course-v1:nalagenetics+PGX101+2020_Q1

### Introduction

Slides 1-3: Introduction of training, speaker and learning objectives

Slide 4: Current healthcare landscape

Slides 5-6: Introducing Nalagenetics

Slide 7: Content Outline

#### Why do Pharmacogenomics Testing?

Slide 8: Title slide

Slides 9-11: Strategies to reduce ADRs

Slide 10: Benefits of PGx testing

Slides 13-17: Reactive vs pre-emptive genotyping

Slides 18-22: Benefits of pre-emptive genotyping

Slides 23-26: Patient case scenario

#### What Fundamentals of Pharmacogenomics I

Slides 27-28: Title slide; Introduction of speaker

Slides 29-31: PGx background

Slides 32-38: PGx terms and definitions

Slides 39-41: Patient case scenario

#### Where and How Fundamentals of Pharmacogenomics I: PGx resources

Slide 42: Title slide

Slide 43: Overview

Slides 44-45: PharmGKB

Slides 46-48: CPIC

Slides 49-51: DPWG

Slide 52: Regulations

Slide 53: Scientific Evidence

Slides 54-57: Patient case scenario

#### Who, When and How Important Pharmacogenes and How to Interpret Them

Slide 58: Title slide

Slide 59: Overview

Slides 60-63: TCAs: amitriptyline and nortriptyline

Slides 64-67: Allopurinol and Steven Johnson Syndrome

Slides 68-73: Codeine

Slides 74-78: Clopidogrel

#### Where and How How do I implement PGx into my Routine Clinical Practice?

Slide 79: Title slide

Slides 80-83: Patient case scenario: conclusion

Slide 84: Nalagenetics workflow

Slides 85-90: Nalagenetics user interface

Slides 91-92: Conclusion

